# Implementing Integrated Genomic Risk Assessments for Breast Cancer: Lessons Learned from the eMERGE Study

**DOI:** 10.1101/2025.05.22.25328180

**Authors:** Cong Liu, Katherine Crew, Jennifer Morse, Jodell E. Linder, Antonis C. Antoniou, Tim Carver, Josh Cortopassi, Josh F. Peterson, Casey N. Ta, Christin Hoell, Cynthia Prows, Eimear E Kenny, Emily Miller, Emma Perez, Gail P. Jarvik, Harris T. Bland, Jacqueline A Odgis, Kathleen F. Mittendorf, Katherine E Bonini, Kyle McGuffin, Leah C. Kottyan, Mary Maradik, Nita Limdi, Noura S Abul-Husn, Priya N Marathe, Sabrina A Suckiel, Sienna Aguilar, Toni J. Lewis, Wei-Qi Wei, Yuan Luo, Robert R. Freimuth, Hakon Hakonarson, Chunhua Weng, Wendy K. Chung, Georgia L. Wiesner

## Abstract

**Objective:** To develop and implement a pipeline for integrated breast cancer risk assessment using the BOADICEA model within the eMERGE study, incorporating polygenic risk scores (PRS), monogenic variants, family history, and clinical factors.

**Materials and Methods:** A pipeline was deployed across ten eMERGE clinical sites, integrating data from REDCap surveys, PRS reports, monogenic reports, and pedigrees via CanRisk Application Programming Interface (API). The process included design, customization, technical implementation, testing, and refinement.

**Results:** The pipeline successfully generated integrated risk scores for >10,000 females. Of these, 3.6% were classified as high-risk (≥25% lifetime risk), and 0.9% harbored rare pathogenic variants in BRCA1, BRCA2, PALB2, or PTEN. High PRS only scores were identified in 5.6% of participants. Among those with high PRS, 34% also had high-risk based on integrated scores. API and User Interface (UI) results were highly concordant, with an average difference of 0.13%.

**Discussion:** Key challenges included integrating diverse data sources, handling missing data, and standardizing pedigree formats. Risk classification discrepancies highlighted the need for refined communication strategies.

**Conclusion:** This study demonstrates the feasibility of PRS-integrated breast cancer risk assessment in clinical settings but underscores challenges in data integration and risk communication. Future work should enhance recalibration for diverse populations and streamline workflows for risk interpretation and update.

## Introduction

Recently, polygenic risk scores (PRS), which reflect the combined effect of numerous common genetic variants identified through large GWAS studies, have been utilized to assess risk for common diseases. While not yet recommended for clinical use^1^, several studies have demonstrated the predictive power of PRS for breast cancer risk in cohort analyses^2–5^. Consequently, the latest versions of breast cancer risk prediction models like Breast and Ovarian Analysis of Disease Incidence and Carrier Estimation Algorithm BOADICEA^6^ (implemented in the CanRisk tool^7^) and Tyrer-Cuzick have incorporated PRS into their risk calculation. Several studies have demonstrated the clinical validity of integrated scores using cohort analyses, particularly when incorporating PRS313^8–14^. However, much of the existing literature emphasizes clinical validity through data analysis, with limited focus on these models’ operations in clinical care.

Although prediction tools with enhanced utility to support online clinical use through innovations like Web Application Programming Interfaces (APIs) and user interfaces have been developed, have been developed to support clinical use, a significant challenge in implementing these tools in real-world settings is that the required variables for assessment are often sourced from multiple platforms and variously structured. For instance, genetic data are typically generated in genetic testing laboratories, either from commercial or academic institutions, and may or may not be uploaded to the electronic health record (EHR). Even when uploaded to the EHR, they’re usually PDFs, limiting direct use of discrete variant data for PRS calculation. Pipelines to collect and automate risk calculation from data gathered across various sources are lacking.

We review the development and implementation of a pipeline for generating an integrated risk score for breast cancer for the ten clinical sites of the eMERGE Network. The pipeline incorporates multiple risk factors that are challenging for clinicians to assess, including monogenic (i.e., rare pathogenic variants in cancer-susceptibility genes) and polygenic risks, family history, lifestyle, and reproductive risk factors into the BOADICEA algorithm^6,15^. It was subsequently deployed across the network, generating and returning breast cancer risk scores to over 13,000 females of diverse ancestry. We detail the involvement of stakeholders, the decision-making processes, and the challenges encountered throughout various stages of implementation. We highlight key “lessons learned” from the eMERGE experience with breast cancer to support the adoption of integrated risk prediction within personalized medicine frameworks in health systems.

## Methods

### Overview of the stakeholders and the implementation work

The primary goal was to create a risk assessment pipeline for breast cancer that can be used across all eMERGE sites^16^. **Figure 1** shows the pipeline development process, which was divided into six stages — design, customization, technical implementation, testing, refinement, and finalization. The stakeholders and their corresponding roles in this process were summarized in **Table 1**. The eMERGE Steering Committee (PISC) leadership established the Breast Cancer Working Group (BCWG) to develop specific protocols for the integration of the breast cancer condition into the overall project. The BCWG proposal, which was reviewed and approved by the PISC, defined the set point for distinguishing “high risk” from “not high risk” and the recommendations for collecting model inputs and generating the integrated breast cancer risk score. The final integrated score was returned to female participants aged 18 and older at enrollment as a component of the eMERGE risk report called the genome-informed risk assessment (GIRA)^16^. The CanRisk Tool^7,17^ (CanRisk.org), which provides access to the BOADICEA model via an API, was selected to generate integrated breast cancer risk scores. It was chosen for its ability to incorporate multiple variables – including PRS – and to accommodate missing data.

**Figure 1.**
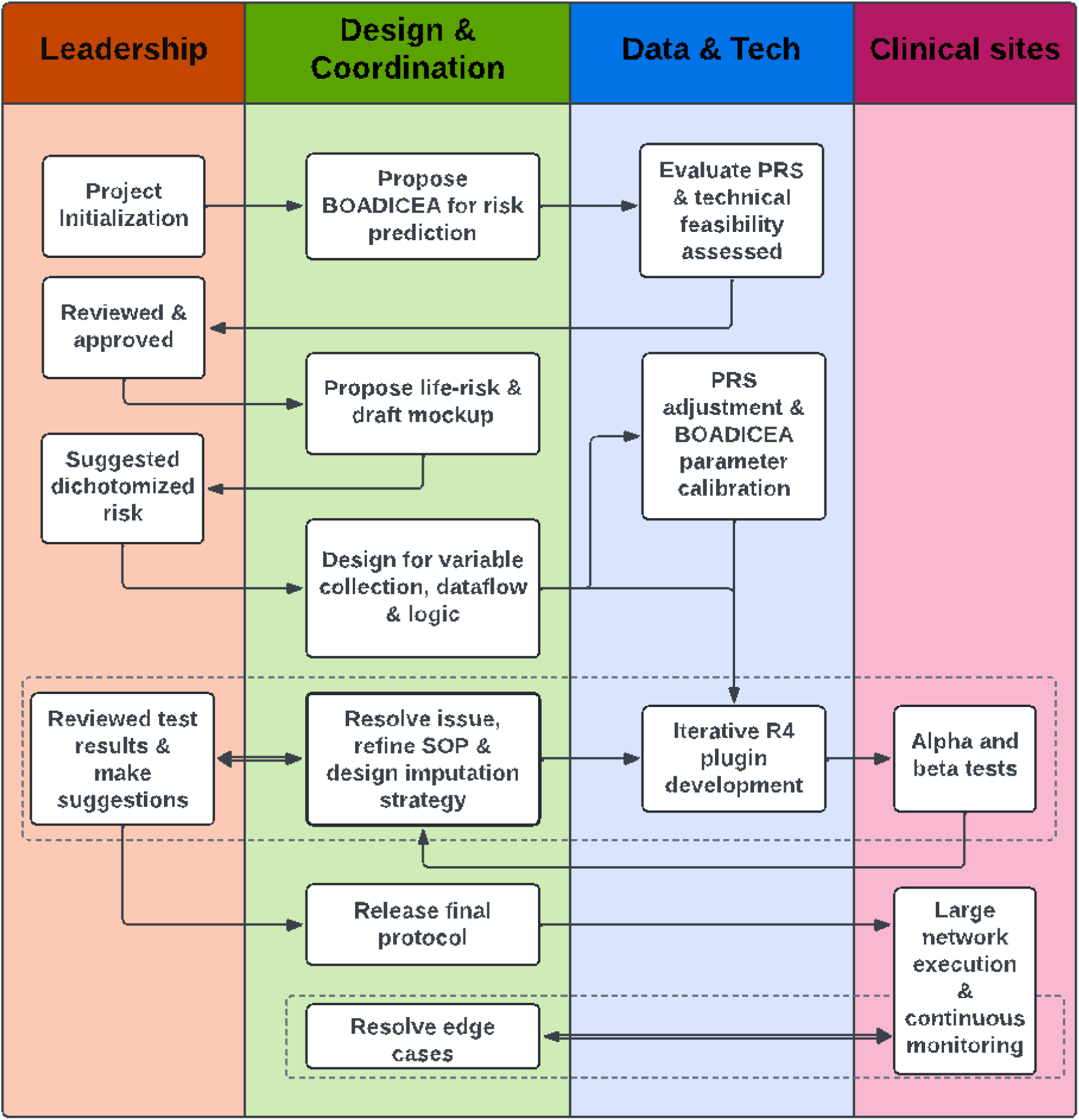
Swanlane diagram illustrating the overview of stakeholders and implementation activities. Different colors/lanes represent distinct stakeholders. Each box indicates a specific task involved in the overall implementation process. Dashed boxes denote iterative processes.

**Table 1.** Description of each stakeholder and their roles.

| **Teams** | **Role** | **Description** |
| --- | --- | --- |
| eMERGE PIs and Steering Committee (PISC) | Leadership | Final decision-makers for the overall eMERGE study |
| Breast Cancer Working Group (BCWG) | Design | Geneticists, oncologists, and informaticians co-lead the project. |
| Coordinating Center (CC) | Coordination | Coordinates all efforts for REDCap survey design, and supports project management, |
| REDCap Team (REDCap) at CC | Technical & Data | Develops R4 extensions and implemented the database design |
| BOADICEA Team (BOADICEA) | Technical | Statisticians built and evaluated the model; Engineers developed BOADICEA web services |
| BROAD Team (BROAD) | Technical & Data | Statisticians built customized PRS models; data engineers deployed the PRS to R4 |
| Invitae Team (Invitae) | Data | Generates monogenic test results |
| MeTree Team (MeTree) | Data | Collects pedigree data |
| eMERGE Clinical Sites (CS) | Clinical | Tests the systems and provides feedback |

The CanRisk team adapted the BOADICEA model to align with the eMERGE study and data inputs. Relevant variables were subsequently collected from various sources, including participant surveys (collected via REDCap^18^), PRS reports (generated by the Broad Institute), monogenic reports (generated by Invitae), clinical data, and pedigrees (collected through MeTree^19^), all centrally stored in a customized REDCap database (R^4^: Recruitment, Results reporting, and Risk Reduction) managed by the coordinating center (CC). Details about the R^4^ implementation and GIRA generation in the eMERGE network have been reported^16^. In the technical implementation stage, the REDCap technical team developed a rule-based mapping program to format the collected data for CanRisk, including converting pedigrees from MeTree to discrete elements. A REDCap plugin was also developed to aggregate and transform all necessary data and automatically trigger the CanRisk API.

For testing, clinical site research staff could access R^4^ to manually initiate or reinitiate the CanRisk score calculation, either through the API or via a user interface where clinical factors, PRS scores, and pedigrees could be entered. Bugs and edge cases (described in the below section) identified during testing were iteratively addressed. Finally, the BCWG developed and released guidelines to properly return the GIRA results to the eMERGE participants at ten clinical sites recruiting adult participants.

### Design: Purpose of the BOADICEA model

eMERGE was originally designed to evaluate the clinical impact of PRS across ten conditions^16^. The BCWG proposed using the BOADICEA algorithm to estimate lifetime breast cancer risk because it integrates monogenic factors (e.g., *BRCA1*, *BRCA2*, *PALB2*), family history, and PRS, which are already used in clinical practice. Versions 5 and 6 of the BOADICEA model include a PRS model with 313 single nucleotide polymorphisms (SNPs) into the risk calculations^12,18^ and the model has been extensively validated^11,13,14,21^.

One limitation is that BOADICEA is primarily based on individuals of European ancestry. Given the challenge of obtaining a comprehensive dataset with all variables across different genetic ancestries, BCWG assessed the validity of the PRS model utilized by BOADICEA (i.e., SNP-313) across diverse ancestries using a previous eMERGE dataset, consistent with other conditions as suggested by the PISC. The results indicated that the SNP-313 model demonstrated reasonable generalizability across four populations: White, Black, Asian, and Latinx^2^. After reviewing these findings, the BCWG’s proposal was approved by the PISC, with a recommendation to develop a pipeline to align PRS scores across different ancestral populations before using the default BOADICEA model to mitigate potential performance disparities across different ancestral populations.

Meanwhile, the PISC and Invitae finalized a sequencing gene panel that mainly includes the Centers for Disease Control and Prevention (CDC) Tier 1 cancer conditions, along with some additional monogenic cancer risks. Three genes—*BRCA1, BRCA2*, and *PALB2*—integral to the BOADICEA (version 5) modeling, were included. A 25% lifetime risk (calculated from age 18 to age 80) threshold to generate a categorical risk (high risk vs. not high risk) rather than a quantitative risk score was selected by the PISC to align with other conditions. This threshold of 25% was chosen to align with current MRI screening guidelines^22^, and to yield a manageable number of high-risk breast cancer participants eMERGE could practically support. We estimated that approximately 2.5-5% of participants would exceed the 25% lifetime risk threshold^2^, which corresponds to about 50-100 females (based on a target of ∼2,500 participants per site and assuming a higher female recruitment rate), who would require high-risk return protocols and care recommendations. This was a feasible number given the budget, person power, and study timeline.

Finally, a GIRA template (**Supplementary Material 1, 2 & 3**) with explanatory language was developed iteratively by CC and BCWG, and subsequently reviewed and approved by the PISC. In the GIRA report, the BCWG decided to separate and explicitly present the monogenic-triggered high risk apart from BOADICEA-integrated risk to ensure that pathogenic/likely pathogenic variants identified in *BRCA1, BRCA2, and PALB2* would receive appropriate attention given the established clinical care guidelines and high risk conferred by variants in these genes.

### Customization: BOADICEA PRS customization

The SNP array utilized in eMERGE comprised only 307 of the original 313 SNPs in the BOADICEA model^6,20^. BCWG recovered a missing SNP (chr6:151955914:A:G) from the original model by substituting it with a surrogate SNP (chr6:151958612:G:T; linkage disequilibrium r > 0.8 based on 1000G), resulting in a final PRS model comprised of 308 SNPs. The BOADICEA team then recalibrated the parameter alpha, an effective size estimation proportional to the odds ratio (OR), based on the Breast Cancer Association Consortium (BCAC) collected samples, which mainly consist of an European population^23^.

Following recommendations from the PISC and Broad team, PRSs for all conditions evaluated in the eMERGE network were recalibrated using *All of Us Research Program* (*AoURP*) samples with genetic ancestry-specific adjustments^24^. This recalibration process was also applied to the breast cancer PRS. While adjusting for non-European populations is still an active research issue, the PISC decided to move forward with an universal OR in this eMERGE network study. Consequently, both the SNP-size adjusted alpha (effective-size) and ancestry-adjusted z-score were documented in the header of the CanRisk input file, available for API invocation or manual adjustments within the user interface (detailed below in **Testing**).

To use the BOADICEA web service for risk calculation, the R^4^ platform hosted at the CC collected data about consented participants, including PRS data from the Broad, monogenic results from Invitae, and pedigrees from MeTree (detailed below in **Technical Implementation)**. These data were aggregated and transformed into a BOADICEA-compatible format and submitted to the BOADICEA web API hosted by CanRisk. The CC established a data-sharing agreement with the CanRisk team to enable the batch processing of research and clinical data by the CanRisk web server.

### Technical Implementation: REDCap/R^4^ extension for BOADICEA calculation

BOADICEA requires a list of clinical and lifestyle factors to assess breast cancer risk (**Table 2**). In eMERGE, a centralized survey platform (i.e., REDCap) was used to collect risk factor data. Since the eMERGE survey was designed to gather information on not only breast cancer-related factors but also risk factors relevant to other conditions, the survey questions were proposed across all conditions, including those focused on breast cancer, and then consolidated centrally to minimize redundancy. As a result, some survey questions did not precisely align with the input requirements of the BOADICEA model. For example, BOADICEA requires height in centimeters, but the R^4^ survey question asks, ‘What is your current height (feet and inches)?’ Additionally, the BCWG decided to exclude the mammographic density question from the survey based on the assumption that most women would not know this information, and would lead to a high likelihood of missing or incorrect data for this question^25^.

**Table 2.** Description of variables and its sources for BOADICEA risk score calculation.

| **Variable** | **Description** | **Source** |
| --- | --- | --- |
| Menarche | Age at menarche | REDCap survey |
| Parity* | Parity | REDCap survey |
| First live birth* | Age at first live birth | REDCap survey |
| OC use | Use of oral contraception | REDCap survey |
| MHT use | Use of menopause hormone therapy | REDCap survey |
| BMI** | Body mass index | REDCap survey |
| Alcohol | Daily alcohol intake in grams per day | REDCap survey |
| Menopause | Age at menopause | REDCap survey |
| BIRADS*** | Mammographic density measured by BI-RADS | Not included |
| Height | Height in cm | REDCap survey |
| PRS BC | Polygenic Risk Score (Breast Cancer) | Broad adjusted PRS z-score and alpha |
| Family history*,** | Family pedigree for calculation | MeTree |
| Monogenic test results | Test results for BRCA1, BRCA2, and PALB2 | Invitae report |

\* Imputed if missing.
\*\* Required variables;
\*\*\* Not collected in this study

A similar challenge arose in providing pedigree data to BOADICEA in the linkage format for defining pedigrees, while eMERGE has partnered with MeTree to collect pedigree information using this software. To address these discrepancies and automate the integrated risk calculation via the BOADICEA web service API, the CC worked with the REDCap Development team to develop a REDCap extension to map the general-purpose survey and MeTree pedigree data into the format required by the BOADICEA API input. The MeTree pedigree includes relatives with unknown medical histories, which could lower the BOADICEA predicted risk (assuming they are healthy). Therefore, our converted CanRisk pedigree data format excluded those individuals. The source code was published in a GitHub repository (https://github.com/vanderbilt-redcap/boadicea-canrisk), and BCWG subsequently reviewed the code to ensure the correctness of the mapping logic.

BOADICEA allows for missing data when estimating lifetime risk. However, to ensure minimal data quality checks, the CC and BCWG decided that the CanRisk API calculation would require only the participant’s sex, height and weight, allowing other variables to be missing. During internal software testing, additional technical requirements were identified in the CanRisk input format. For example, CanRisk does not allow missing age data in the pedigree for affected individuals. To make full use of family history information, BCWG proposed an imputation strategy for missing ages of family members, guided by predefined rules. To stay on track with the overall timeline, the imputation strategy was developed somewhat arbitrarily without throughput validation, aiming for the most reasonable assumptions possible. Specifically, for each generation removed from a target node, missing ages were imputed by adding or subtracting 25 years based on the generation of the relationship. Additionally, ages were imputed based on diagnosed age when available, and conversely, diagnosed age was imputed based on current age. In addition, some sites reported high missingness in completing the MeTree pedigree due to the complexities of data entry. To address this issue, the BCWG proposed and implemented a secondary survey, which required minimal family history information to be used in constructing the CanRisk pedigree. Additional technical issues were discovered during testing, leading to more imputations to ensure the GIRA report included as many participants as possible.

### Testing & Refinement: Compare UI and API generated risks

An alpha version test was then conducted to ensure the correctness of the risk calculation in production by comparing the API-returned risk assessments (“API”) with results manually inputted by the clinical research staff at two clinical sites using the CanRisk user interface (“UI”). To ensure consistency between API input and UI input, multiple QC training sessions were held to instruct site research staff on how to use the R^4^ data and apply the same imputation strategies if needed. Additionally, staff at each eMERGE site were trained to manually enter the data into the CanRisk UI. During this test phase, the CC maintained error logs to document discrepant cases (subjectively identified by site staff). The clinical research staff from two testing sites, along with the BCWG and the CC, discussed these cases to address potential issues. Through this process, we promptly identified and resolved numerous issues (**Table 3**). Some issues required changes to the original code, so the BCWG submitted code change requests through GitHub, which were reviewed and merged by the REDCap development team.

**Table 3.** Issues identified in the risk score generation, review and return process.

| **Issue** | **Details** | **Examples** | **Solution** |
| --- | --- | --- | --- |
| Technical Issues | Code bugs or API downtime | Encoding error of oral contraceptive use; BOADICEA API is down. | Fix the code. |
| Input Issues (a) - Data Error | Incorrect data entry or logical errors in the input which can stop BOADICEA generate a prediction | Age is not an integer; Condition age in MeTree is larger than current age; Males with ovarian cancer in MeTree; Females with prostate cancer in MeTree. | Add data validation and purity checks in the code. |
| Input Issues (b) - Data Missing | Missing critical information in the input which causes BOADICEA to ignore the useful information or make wrong assumptions | Ages of relatives or ages of conditions are missing in the pedigree. Relatives with medical history unknown will be treated as healthy in BOADICEA calculation. | Implement imputation guidelines to fill missing data and exclude relatives with unknown medical history. |
| Input Issues (c) - Data Discrepancies | Conflicting data between sources or fields | Different ages in the survey and MeTree entry; Parity entry in the survey differs from the number of children in MeTree. | Implement guidelines to resolve data conflicts. |
| Interpretation Inconsistencies | Input from research staff in Canrisk (UI) does not match R4 extension generated input | Research staff used a different imputation approach when a relative's age was missing, causing discrepancies between UI input and R4 extension-generated API input. | Conduct training sessions and establish a standard protocol. |
| Edge Cases | Rare or unexpected situations causing errors | MeTree formatting of pedigree data for twins was causing an age-related error. | Research staff review edge cases and resolve them on a case-by-case basis. |

After running the alpha test for two months (from December 2022-January 2023), and addressing issues encountered, the network advanced to a beta version test by including more clinical sites. In the beta test, we focused our review on cases for whom the API-generated lifetime risk exceeded 20%, comparing these with the UI-generated (by site research staff) risks. To minimize discrepancies caused by individual interpretations of clinical variables or pedigree differences, the CC re-ran the UI risk assessments. If discrepancies persisted, the BCWG further investigated these cases, leading to the identification and resolution of additional issues during this phase.

### Finalization: Release Guidelines for Score Generation, Review and Return

After the beta test, the BCWG established guidelines to address risks at the border of the threshold for high-risk and non-high-risk categorization (e.g., 25%). Site research staff were instructed to manually review cases when the breast cancer risk was between 20-25%, based on automated CanRisk API calculations, while an independent score was generated using the CanRisk UI. Cases above 25% were excluded from manual comparison, as our review of the alpha and beta test results indicated that these cases are highly likely to be high risk. Subsequently, we applied a set of rules to borderline cases (20-25%) determine the final status: (1) If the UI score was below 25%, the case was classified as not at study threshold for high risk; (2) If the UI score is equal to or greater than 25%, BCWG reviewed the case together with the site staff to make the final determination. Finally, the entire risk generation, review, validation and return process (**Figure 2**) is composed of multiple phases and includes five checkpoints.

**Figure 2.**
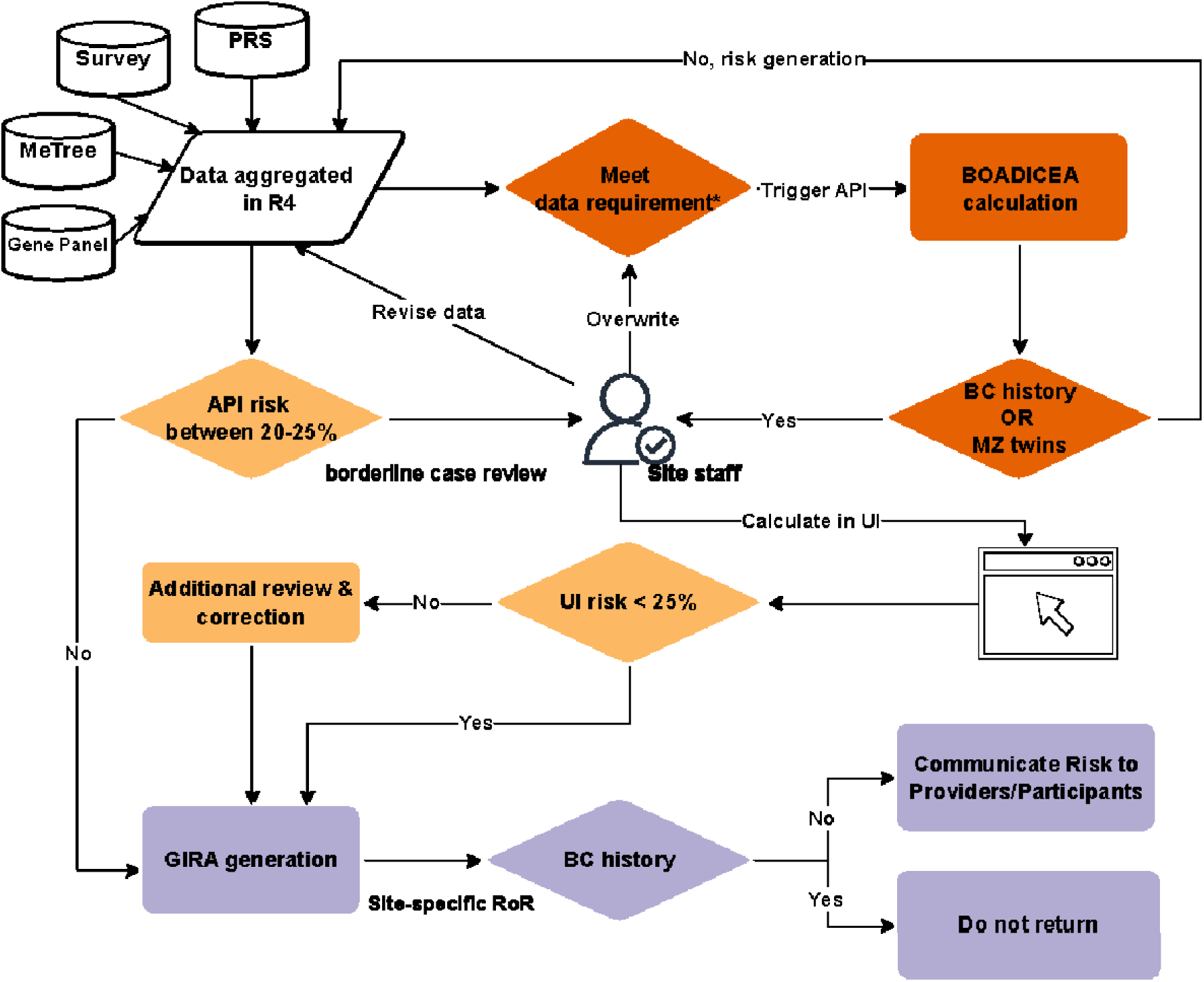
Finalized standard operating procedure (SOP) for the risk generation, review, validation, and return process. The entire procedure is composed of multiple phases (different colors) and includes five checkpoints (diamond-shaped boxes). **Phase 1**: Before API query, the participant must have sex assigned as female at birth, 18 years and old, and provided baseline height and weight. If MeTree data are unavailable, the site’s research staff acknowledged its absence, and in such cases, the R^4^ extension generated a placeholder pedigree containing only healthy parents (father and mother). Other missing data (e.g. PRS) were permissible. For a successful API risk generation, the participant must have no prior breast cancer history recorded in the MeTree, and no monozygotic twins; **Phase 2:** Borderline cases were review. Sites then verified that data from Broad, Invitae, MeTree, and other sources was reviewed and approved before generating the GIRA report, which includes conditions beyond breast cancer risk. **Phase 3**: Sites implemented their own high-risk return protocols to communicate risk to participants and their healthcare providers. If additional information (e.g., from the EHR) revealed a prior history of breast cancer, sites were instructed not to return a high-risk breast cancer result.

## Results

### High risks in different categories

As of October 2024, this pipeline generated and returned integrated BOADICEA scores for 10,928 eligible females, all aged 18 and older, who consented and had BMI data. Among these women, 398 (3.6%) were identified as having an integrated lifetime breast cancer risk of 25% or higher, classifying them as high-risk according to the study design. When assessing the participants with elevated risk using PRS without additional variables (PRS only), 608 women (5.6%) were classified as ‘high PRS risk’ based on their adjusted PRS score. This ratio aligns well with the definition of ‘high PRS risk,’ where the adjusted PRS exceeds the top 5% threshold in the *AoURP* reference panel. Among those classified as high risk by PRS alone, 207 women (34.0%) also had a high integrated risk, a 9.4-fold increase in the high risk ratio compared to the reference group (3.6%). The mean integrated risk for the high PRS only group was 23.4% (std: 7.5), compared to 11.8% (std: 6.6) in the non-high PRS group. A breakdown of the risk distributions in major self-reported race/ethnicity groups in **Table 4**.

**Table 4.**
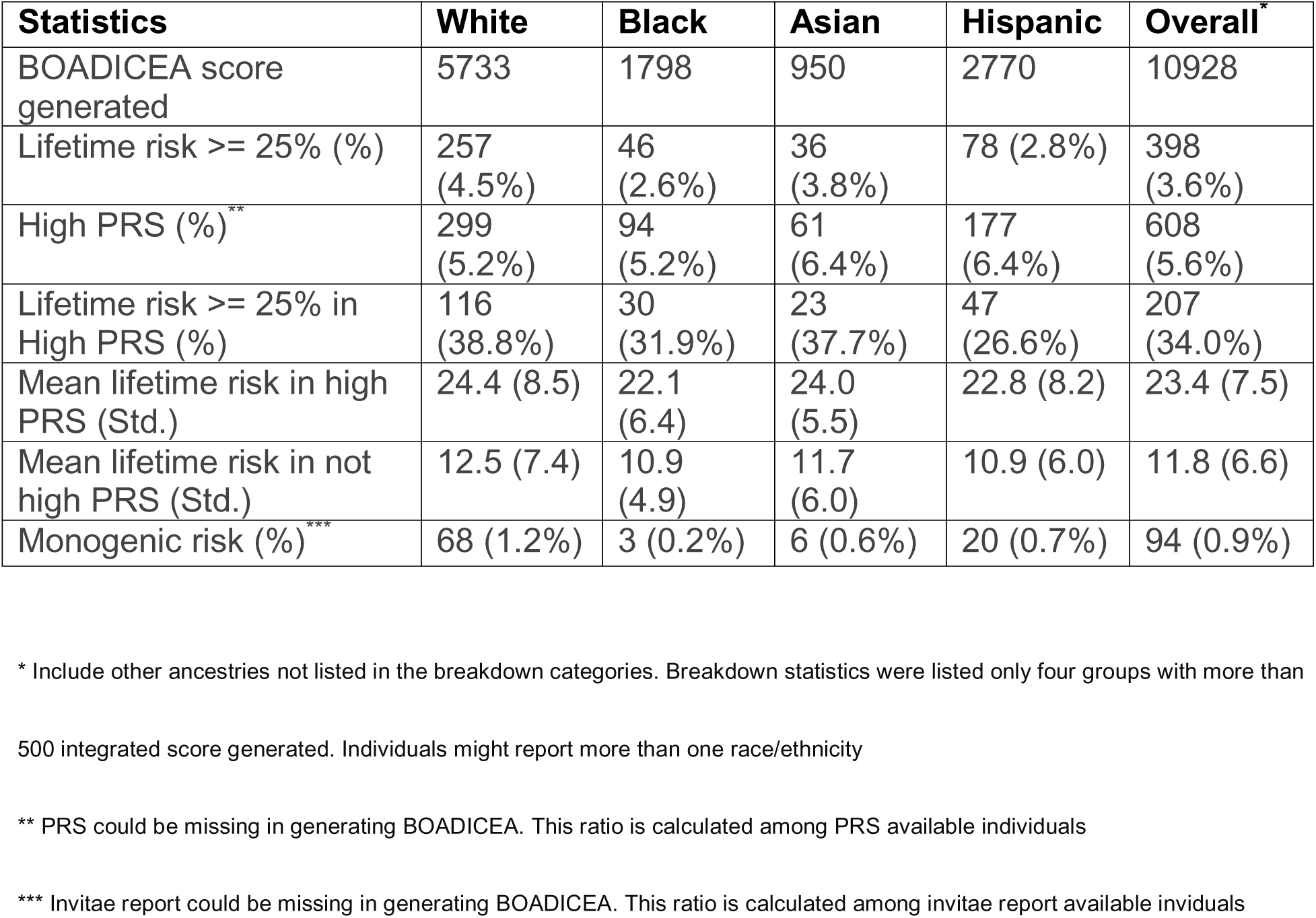
Distribution of life-time risk and PRS in different self-reported race/ethnicity groups.

| **Statistics** | **White** | **Black** | **Asian** | **Hispanic** | **Overall\*** |
| --- | --- | --- | --- | --- | --- |
| BOADICEA score generated | 5733 | 1798 | 950 | 2770 | 10928 |
| Lifetime risk ≥ 25% (%) | 257<br>(4.5%) | 46<br>(2.6%) | 36<br>(3.8%) | 78 (2.8%) | 398<br>(3.6%) |
| High PRS (%)** | 299<br>(5.2%) | 94<br>(5.2%) | 61<br>(6.4%) | 177<br>(6.4%) | 608<br>(5.6%) |
| Lifetime risk ≥ 25% in High PRS (%) | 116<br>(38.8%) | 30<br>(31.9%) | 23<br>(37.7%) | 47<br>(26.6%) | 207<br>(34.0%) |
| Mean lifetime risk in high PRS (Std.) | 24.4 (8.5) | 22.1<br>(6.4) | 24.0<br>(5.5) | 22.8 (8.2) | 23.4 (7.5) |
| Mean lifetime risk in not high PRS (Std.) | 12.5 (7.4) | 10.9<br>(4.9) | 11.7<br>(6.0) | 10.9 (6.0) | 11.8 (6.6) |
| Monogenic risk (%)*** | 68 (1.2%) | 3 (0.2%) | 6 (0.6%) | 20 (0.7%) | 94 (0.9%) |

\* Include other ancestries not listed in the breakdown categories. Breakdown statistics were listed only four groups with more than
500 integrated score generated. Individuals might report more than one race/ethnicity
\*\* PRS could be missing in generating BOADICEA. This ratio is calculated among PRS available individuals
\*\*\* Invitae report could be missing in generating BOADICEA. This ratio is calculated among invitae report available individuals

While monogenic risks had a separated return workflow, we examined their integrated risk retrospectively. There were 94 women (0.9% of 11,577) found to carry rare pathogenic variants (PV), including *BRCA1* (25), *BRCA2* (47), *PALB2* (22), and *PTEN* (2). Two participants with a *PTEN* PV had low integrated risk, as PTEN PVs are not considered in BOADICEA. All others have high risk based on integrated scores.

### UI vs API generated results

At one research site (UAB), a quality control project was undertaken to compare risk scores generated by the CanRisk API with those produced through its web UI. Risk scores for 733 participants, initially calculated using R^4^ via the API, were manually recalculated through the online tool. This process involved thoroughly reviewing, interpreting, converting, and inputting all available patient data, including proband pedigrees and survey responses from multiple instruments, into the UI. The average lifetime risk scores from the UI closely aligned with those from the API, showing an average difference of just 0.13% (**Table 5**). Most discrepancies were attributed to variations in the number of data points entered and potential imputation inconsistencies. Overall, the API and UI scores exhibited strong concordance within this subset of eMERGE participants.

**Table 5.** Comparison of risk estimates generated via API and UI approaches.

| **Methods** | **Average lifetime risk score** | **Lifetime Risk Score Range** | **Medium Lifetime Risk Score** |
| --- | --- | --- | --- |
| API<sup>*</sup> | 11.11 | 2.8 - 90.8 | 9.9 |
| UI<sup>**</sup> | 11.24 | 2.4 - 90.5 | 9.9 |

**\*API** (Application Programming Interface): Risk scores were generated automatically using a REDCap plugin to collect, format, and submit input variables to the CanRisk API.
**\*\*UI** (User Interface): Risk scores were generated manually by reviewing collected data and entering variables into the CanRisk web interface to run the calculation.

### Issues encountered during multiple testing phases

Over multiple testing phases, site coordinators entered 174 questions and potential errors in a log for review. The coordinating center staff reviewed and triaged these issues and worked with members of the BCWG for adjudication. The major issues we encountered in the two testing phases are summarized in **Table 3**. We categorized the issues into four main groups: technical issues, input issues: interpretation inconsistencies: and edge cases.

### Discrepancy between MeTree Pedigree and CanRisk data format

One of the key lessons we learned from this implementation study was the need to develop a standardized pedigree data format that allows pedigree data collected by MeTree to be used as input for other software (CanRisk). Creating pedigrees can be complex for participants and time-consuming, so reusing them for various risk calculations requires a standardized data adapter to connect pedigree programs.

Among all the issues above, many of them were caused due to the incompatibility of the MeTree pedigree and the pedigree data required by CanRisk. **Table 6** provides a summary of the major differences. Minor differences, such as ‘female’ versus ‘F’ or 16-digit ID encoding versus 7-digit ID encoding, are not listed. As a result, a complex mapping schema was developed and implemented in the eMERGE specific REDCap plugin to resolve the conflicts and, in some cases, perform imputations. The source code is available at https://github.com/vanderbilt-redcap/boadicea-canrisk.

**Table 6.** Pedigree representation differences between MeTree and CanRisk.

| **Gaps** | **MeTree** | **CanRisk** |
| --- | --- | --- |
| Overall Structure | JSON array, with each element representing a relative. | Embedded in a general input, with headers indicating lifestyle and PRS risks, followed by each row represents a relative. |
| Relationship | Direct relationship between the target and relatives is coded (e.g., MGRMTH for maternal grandmother). The relation 'SELF' indicates the participant. | Each row contains parent IDs representing an edge in the pedigree, with a column called "target set 1" indicating the participant. |
| Missing Tolerance | Allows situations where only the father or mother node is present for an entity; current age can be missing. | A complete pedigree is required. If the current age is missing, the node is dropped. If the age is '0' or outside the range of 1-125, the node will also be dropped. |
| Age Allowance | Digitized numbers allowed; The condition age can be older than the current age. | Age must be an integer; Strict age check |
| Multiple Diagnoses of Cancers | Multiple diagnoses must be inferred from age at diagnosis; more than two occurrences are possible but may not indicate bilateral diagnosis. | Only two diagnoses are allowed for breast cancer, indicating bilateral diagnosis. |
| Gender Restriction | No gender-related condition checks. | Females cannot have prostate cancer, and males cannot have ovarian cancer. |
| Medical History | Unknown' is allowed. Healthy is specifically indicated to represent healthy individuals | If no condition is provided for a node, it is assumed to be healthy. |
| Genetic Testing | Genetic variant status is not documented. | Can input genetic testing results for various cancer-related variants. |
| Conditions Documented | A wide range of conditions is documented, with more general categories. | Only cancer types, including breast, prostate, ovarian, and pancreatic, are documented. |

## Discussion

While other studies have evaluated BOADICEA with integrated PRS in predicting breast cancer risks^16^, this is the first study aimed at operationalising BOADICEA multifactorial risk assessment within clinical studies in multiple institutions. The study has several unique features. First, the study population is diverse, with more than 50% of individuals self-reporting non-European races and ethnicities. This required adjustment to the original PRS, which was developed based on European populations. Second, the integrated score includes information on rare pathogenic variants in cancer susceptibility genes derived from a targeted gene sequencing panel, reflecting a more realistic clinical setting, as gene-panel testing is currently more widely applied than PRS. Third, the genotype data used in eMERGE were designed to calculate multiple disease PRSs, not only for breast cancer, which could represent a more cost-effective approach for PRS-integrated risk assessment in clinical practice. As a result, the PRS used in this study differs from that in the original risk prediction model, necessitating recalibration of the BOADICEA parameters for the PRS effective size. Lastly, this study also incorporates the return of results, with a standardized protocol to generate reports and communicate risk status to individuals based on their specific risk level and follow the participants to understand impact on clinical care.

To fully understand the actions and decision-making processes within this breast cancer-specific workflow, there are two key points. First, though presented linearly, work often occurred concurrently in iterative cycles. Second, some decisions, made at a higher level, applied to all conditions in the eMERGE network. We recommend that readers refer to the comprehensive view of eMERGE’s design^16^. This embedded design introduces several challenges in our risk-return process. While the BCWG initially suggested returning continuous risks, the leadership opted for a dichotomized approach, categorizing risk as high or not high, to align with other conditions. This dichotomous classification could have significant implications for borderline cases. While a 1% change in lifetime risk might not impact clinical decisions in practice, in a dichotomized scenario, it could mean the difference between being classified as high risk (25.5%) or not high risk (24.5%). Unlike monogenic result returns, where the boundaries are more clearly defined, integrated risks are quantitative and continuous. In clinical practice, quantitative risk information may be preferred or, at the very least, should be provided alongside categorical classifications to offer a more complete context for decision-making. In addition, most conditions in the eMERGE study do not have an established risk model like breast cancer, which integrates PRS, monogenic data, family history, and other clinical factors. The risk classification for most other conditions was based on PRS alone. However, when multiple types of reports—such as monogenic reports, PRS reports, and integrated reports—are returned together, confusion can arise because PRS is only one component of breast cancer risk. Although our preliminary analysis showed that PRS had a significant impact on the final risk (9.4 times higher than baseline), a large portion of individuals classified as ‘high risk’ based on PRS alone (>60%) were not classified as ‘high risk’ when integrated risk across all factors was calculated. Looking ahead, we anticipate the development of more comprehensive genomic risk prediction models for many conditions. Until then, risks based on a subset of factors, such as PRS, should be interpreted with caution and within the broader clinical context.

Another challenge was coordinating across multiple systems (e.g., MeTree, REDCap, and CanRisk) and stakeholders within our modular design, which allows different teams to work on separate components. For example, due to differing requirements for data completeness across platforms—where certain variables are optional on one platform but required on another—we adopted an imputation strategy to maximize data utility when required variables were missing. Also, the update of one module could trigger the change of the entire workflow. For example, during the study, the BOADICEA model was upgraded from version 5 to 6^21^, with the UI supporting only version 6. To maintain consistency, we kept using the archived version 5 for risk calculation and reporting, though this complicated comparisons between API and UI-generated risks, as they relied on different versions. This prompted discussions on appropriate reanalysis timing when prediction models are updated. While this issue has been widely discussed in the context of variant reclassification, the influence of PRS and other risk factors on overall risk is not as significant as that of high-penetrance pathogenic variants. Updates may also have differential impact on genetic ancestral groups. As such, a separate discussion and potentially different actions may be warranted compared to variant reclassification. Based on our experience in this study, we anticipate that this issue of differential impact will become increasingly common, particularly with advancements in AI-based predictions. Furthermore, while this study focused on lifetime risk assessment, it is important to recognize that some women may need decade-by-decade risk information to support both immediate and long-term decision-making, as risks and personal priorities evolve with age^26^. Determining when to rerun the risk model and re-communicate results is crucial, requiring input from all stakeholders—including clinicians, patients, and experts in ethics, legal, and social implications (ELSI)—to fully understand the ethical and practical impacts and to develop patient-centered, feasible recommendations.

## Data Availability

All data produced in the present study are available as supplementary materials.

## Author Contribution

**C.L., G.L.W., J.M., C.W., W.K.C., J.F.P., K.D.C., R.R.F., and H.H.** contributed to conceptualization; **C.L., G.P.J., W.-Q.W., N.L., J.C.,** and **T.J.L.** contributed to data curation; **C.L.** contributed to analysis; **C.L., J.E.L., J.M.,** and **K.M.** contributed to methodology; **C.L., G.L.W., J.C.,** and **W.K.C.** contributed to manuscript drafting; **P.N.M., K.E.B.,** and **J.A.O.** contributed to investigation**; H.T.B., G.P.J., J.E.L., C.W.,** and **J.F.P.** contributed to project administration; and **C.H., E.E.K., N.S.A., S.A.S., S.A., M.M., Y.L., E.M., E.P., C.P., L.C.K., K.F.M., C.N.T., T.C.,** and **A.C.A.** contributed to manuscript review/revision.

## Acknowledgments & Funding

The network would like to thank the individuals enrolled in the study, the VUMC data core team, team members at the Broad Institute, Invitae Corporation, Duke University (MeTree), and CanRisk team that helped set up the automated infrastructure for data transfer to R4. This phase of the eMERGE Network was initiated and funded by the NHGRI through the following grants: U01HG011172 (Cincinnati Children’s Hospital Medical Center); U01HG011175 (Children’s Hospital of Philadelphia); U01HG008680 (Columbia University); U01HG011176 (Icahn School of Medicine at Mount Sinai); U01HG008685 (Mass General Brigham); U01HG006379 (Mayo Clinic); U01HG011169 (Northwestern University); U01HG011167 (University of Alabama at Birmingham); U01HG008657 (University of Washington); U01HG011181 (Vanderbilt University Medical Center); U01HG011166 (Vanderbilt University Medical Center serving as the Coordinating Center). The CanRisk API, ACA and TC are supported by Cancer Research UK grant PPRPGM-Nov20\100002 and by core funding from the NIHR Cambridge Biomedical Research Centre (NIHR203312) [*]. *The views expressed are those of the author(s) and not necessarily those of the NIHR or the Department of Health and Social Care.

## Notes

### Competing Interest Statement

The authors have declared no competing interest.

### Funding Statement

This study was funded by National Institute of Health.

### Author Declarations

The central IRB at Vanderbilt University Medical Centre (IRB 211043) gave ethical approval for this work.

