## Supplemental 1 for "Implementing Integrated Genomic Risk Assessments for Breast Cancer: Lessons Learned from the eMERGE Study"

| | |
|---|---|
| Record ID | 115673 |
| GIRA Report Date | 03-17-2025 |
| GIRA Report Date | 2025-03-17 |

#### eMERGE Study Overview

eMERGE is a research network of 10 academic medical centers across the United States. It is funded by the National Human Genome Research Institute (NHGRI). The eMERGE study aims to find better ways to assess and manage patients' risk for future health conditions. eMERGE wants to learn if risk based on genetics, family health history, and personal health history, helps patients and their doctors make health care choices. This study is developing a Genome Informed Risk Assessment (GIRA) report. The GIRA report will have information about genetic risk, clinical risk, and family history for certain common conditions in adults and children.

#### Sections included in the GIRA Report

- Summary of findings
- Participant frequently asked questions
- Study methods and limitations
- Invitae eMERGE panel screen report
- Broad Institute Polygenic Risk Report
- MeTree family health history pedigree

#### Site Contacts


| | |
|---|---|
| Vanderbilt University Medical Center<br>Principal Investigator<br>Dan M. Roden, MD<br>phone: 615-322-0067<br> | Vanderbilt University Medical Center<br>Study Coordinator<br>Harris Bland, MPH, MBA<br>phone: 615-875-8240<br> |


#### Genome Informed Risk Assessment (GIRA)

<table>
<tr><td rowspan="7">The following report is a summary of evaluated risk factors for common conditions. Common gene changes were tested for the conditions listed. Rare gene changes and family history were also included for some conditions. This study determined who is at high risk based on specific criteria for each condition.</td><td colspan="2">Risks Evaluated:</td></tr>
<tr><td>Asthma*</td><td>Hypercholesterolemia</td></tr>
<tr><td>Atrial fibrillation</td><td>Obesity/BMI**</td></tr>
<tr><td>Breast cancer</td><td>Prostate cancer</td></tr>
<tr><td>Chronic kidney disease</td><td>Type 1 diabetes*</td></tr>
<tr><td>Colorectal cancer</td><td>Type 2 diabetes**</td></tr>
<tr><td>Coronary heart disease</td><td></td></tr>
</table>

* children only **adults and children

| | | |
|---|---|---|
| eMerge network ELECTRONIC MEDICAL RECORDS AND GENOMICS | **Name:** Jane Smith | **Provider:** John Tester |
| | **DOB:** 1984-01-01 | **Site ID:** Vanderbilt University Medical Center |
| | **Sex at birth:** Female | **Participant Lab ID:** L712345677 |
| | **Date of Report:** 03-17-2025 | |

#### Summary of Findings

Risk assessment limitation: Family history is a component of overall risk status for some of the conditions in this study (Atrial fibrillation, Breast cancer, Chronic kidney disease, Coronary heart disease, Prostate cancer). Family history is self-reported and risk status may not be accurate if individuals report incomplete or unknown family history.

**RESULT:**

**This participant was not at high risk for any of the conditions evaluated.**

This means the polygenic risk scores (PRS) for these conditions did not meet the threshold for high risk. For some conditions, other factors such as sequencing of specific genes (monogenic risk), or family history, also contributed to the risk estimate. For breast cancer, a high risk PRS alone does not meet the threshold for high risk in this study. In addition, you may not receive a score if you have a personal history of breast cancer. Review the methodologies section for a full explanation of how high risk for each condition was determined.

For participants already diagnosed with a condition, providers should continue with current treatment.

**Please note that a person's overall risk for any of these conditions could still be higher than the general population based on factors that are not included in the GIRA.**

Information contained in this report does not replace evaluation by a health care provider. General risk reducing strategies such as maintaining a healthy lifestyle and age recommended screening tests are still recommended. Questions about what this report means for this individual's medical management should be discussed with their provider.

### Frequently Asked Questions

#### What is the Genome Informed Risk Assessment (GIRA) or health risk report?

The Genome Informed Risk Assessment (GIRA) is the overall report from this research study. This report says if you/your child are at high risk for any of the health conditions studied. “High risk” means that you have a higher chance of getting a condition than the average person. This study looked at 9 conditions for adults and 4 conditions for children under 18.

| Health Conditions Assessed | |
|---|---|
| Asthma (children) | Hypercholesterolemia (adults) |
| Atrial Fibrillation (adults) | Obesity (children and adults) |
| Breast Cancer (adults) | Prostate Cancer (adults) |
| Chronic Kidney Disease (adults) | Type 1 Diabetes (children) |
| Colorectal Cancer (adults) | Type 2 Diabetes (children and adults) |
| Coronary Heart Disease (adults) | |

This report has information about health risks including genetic (one or more genes), you/your child's health history, and family health history. If you were found to be at high risk for one or more conditions, you can learn more about your risk in the “Understanding Your Risk” pages. You can speak to a study staff member about your results. This report may be put in your electronic health record and will be given to your doctor. It contains a summary of your risk factors, as well as the gene reports and family history information described in the consent form.

#### What is monogenic risk?

A single difference in one gene can have a big impact on a person’s risk for developing a health condition. This is called monogenic risk because “mono” means “one.” For adults in this study, your health risk report includes genetic testing results for a small number of monogenic risks. Children in this study were not tested for monogenic risks. You should talk to your doctor about your results from this study. Your doctor may recommend changes in your healthcare.

#### What is a Polygenic Risk Score (PRS)?

Everyone has thousands of genetic differences. Some genetic differences can slightly increase the risk for developing a health condition. A polygenic risk score (PRS) is made by adding up these small genetic risks. It is called polygenic risk because “poly” means “many.” A PRS is used to estimate the overall risk someone has of developing a health condition. PRS is a new method for estimating risk. Scientists are still working to improve PRS for different conditions. This study is trying to see if these risk estimates are helpful for you and your doctor(s). You should talk to your doctor about your results from this study. Your doctor may recommend changes in your healthcare.

#### How should I talk to my doctor about my results?

You should share this report with your doctor. You should ask your doctor what the results mean for your health. You should ask your doctor to help you follow any recommendations made in the report.

#### What are the possible results for each condition?

You can receive two types of results for each condition tested. These results will either be 'high risk' for a condition OR 'not at high risk' for a condition.

- You can be 'high risk' for the following reasons:
    - You had a positive monogenic result. A positive monogenic result means a genetic difference in one of your genes that is known to increase risk was found. OR
    - Your PRS was above the study threshold for a condition. The study "threshold" is the point at which someone's risk for developing a condition is higher than the average person. OR
    - You have one or more close family members with the condition.
- You can be 'not at high risk' for the following reasons:
    - You had a negative monogenic result. A negative monogenic result means no genetic differences that increase risk were found. AND
    - Your PRS was below the study threshold for a condition. The study "threshold" is the point at which someone's risk for developing a condition is higher than the average person. In this study, the researchers set a specific threshold to indicate high risk that is unique to each condition. OR
    - You did not have a strong family history for the condition.

#### How accurate are these results?

The GIRA health risk report gives a summary of many different types of risk factors. The accuracy of your family history risk depends on how much information you knew about your relatives. The accuracy of your clinical risk factors depends on responses to the surveys and information from your electronic health record. Your genetic risk is made up of monogenic and polygenic tests. Monogenic tests have been around for a long time and have clear recommendation guidelines. Your polygenic risk score results are based on what science currently knows about genetic differences that impact a person's risk for these conditions. The GIRA health risk report is <u>not</u> diagnostic. This research study measures *risk (or the chance) of developing a health condition*. Scientists cannot know for sure who will develop a disease and who will not. Health conditions can have multiple causes.

Genetic research studies need a large number of diverse participants to answer questions about our genes. Many polygenic risk scores were developed using data mostly from people of European descent. The study PRSs may not be as good at estimating risk in people who are not of European descent. This study tried to make PRSs that used genetic information from people of many different races, ethnicities, or ancestries. Where possible, the results have been validated (or confirmed) in people from four populations: Asian descent, African descent, European descent, and Hispanic/Latino descent. However, this type of information was not always available for every condition. This may impact how well your results estimate your risk for the health conditions. Please refer to the methods section to learn more about how this limitation may impact your results.

You may identify with more than one, none, or all of the listed populations. Across the different results you receive in your GIRA, some may still be meaningful even if you don't identify with the populations mentioned. You should discuss all of the results you receive with your doctor. Your risk likely falls within the range of risk presented in the report. Your participation in this study may help improve health care for all people in the future.

#### What does it mean if I am at high risk for one or more conditions?

Being at high risk for one or more conditions means you are in the top 2-10% of the population for developing a condition based on your polygenic risk score. This means you have a higher chance of developing that health condition than the average person, but does not mean you will definitely develop the condition. You may be at high risk due to your genetic results or family history. The "Understanding Your Results" pages in this report will help you understand what your high risk results mean for your health and your medical care.

#### What does it mean if I am not high risk?

Being defined by the study as not high risk does not mean you are at low risk for developing that condition. Your overall risk for any of these conditions could still be higher than the average person based on factors that were not looked at in this study.

#### What if I already have one of the conditions studied and I am found to be at high risk?

There are many causes that can lead to developing a health condition. Other aspects of life such as lifestyle, environment, random chance, or other genetic differences not studied may be part of why you were diagnosed with the health condition.

#### What if I already have one of the conditions studied and I am not given a high risk result?

There are many reasons that someone develops a health condition. This research study measures risk from certain causes (such as your genetics and family history). Sometimes people who do not have any risk factors will still develop disease. Lifestyle, environment, random chance, or other genetic differences not studied may be part of why you were diagnosed with your health condition.

##### What do these results mean for my family?

Monogenic risk has been studied for a longer time and we have more information about these genes. We know that your monogenic risk is important information for your blood relatives to know. PRSs are new and are still being studied. We do not currently know what your PRS results mean for your blood relatives. You should talk with your family members and they should discuss your family medical history and your test results with their own doctor(s).

##### Could my results change?

Your genes do not change, but science does. New tools to estimate risk could become available. The interpretation of your genetic differences could change. These research results were generated using the most up to date knowledge and information available at the time this report was written.

##### What other factors might influence my risk that are not accounted for?

This report is based on what we currently know about genetic differences that impact a person's risk for the conditions tested in this study. However, there are many factors that influence risk. Some of these factors are understood by scientists and doctors, but there are many factors that are still unknown. Some factors change over time like a person's age, lifestyle, environment, medications, or diet and their risk for disease can change as well. People can have genetic differences that have a small or large impact on risk. Not all genetic differences that influence risk for disease were tested in this study. Family history and clinical risks factors were evaluated based on the information available. If any of the information used to estimate risk was missing, your risk estimate may not be as good. Children were only tested for a few of the conditions included in this study. A list of the conditions tested can be found on the first page of this report. Please refer to the methods section to help understand how the risk estimates apply to you.

#### About this study

The genome integrated risk assessment (GIRA) was developed as part of a research study by the Electronic Medical Records and Genomic (eMERGE) network and funded through the National Institute of Health (NIH). This study was approved by the central institutional review board at Vanderbilt University Medical Center (IRB 211043).

#### Methods

**Monogenic sequencing:**
Sequencing for monogenic variants in 16 genes was performed by Invitae, a CLIA accredited laboratory. This custom proactive panel test includes the following genes: *BRCA1, BRCA2, MLH1, MSH2, MSH6, PMS2, EPCAM, APOB, LDLR, LDLRAP1, PCSK9, PALB2, PTEN, STK11, TP53, LMNA*. See the Invitae report attached for a description of methods.

**Polygenic risk calculation:**
Genotyping was performed at the Broad Institute using the CLIA accredited Global Diversity Array from Illumina, Inc. Polygenic risk scores for each condition were calculated from the genotyping data. See the attached Broad eMERGE Polygenic Risk Report for a description of methods.

**Family History:**
Family history information was self-reported by the participant or their parent/caregiver using MeTree software developed by the Duke Center for Applied Genomics and Precision Medicine. The participant's pedigree was generated using MeTree.

**Clinical data:**
Clinical data used in this study were obtained from the participant's electronic health record with participant/parent consent or from self-report via participant/parent/caregiver completed surveys. Clinical data, along with genetic data, was used to estimate risks for breast cancer and coronary heart disease as part of integrated risk scores. Clinical data for other conditions may be displayed for provider reference.

**Care Recommendations:**
Care recommendations were generated by members of the eMERGE network and represent the collective recommendations of disease experts, clinical care providers, and researchers.

**Integrated Scores:**
Breast cancer: Breast and Ovarian Analysis of Disease Incidence and Carrier Estimation Algorithm (BOADICEA) (https://ccge.medschl.cam.ac.uk/boadicea/) predicts breast cancer risk based upon the family history, lifestyle/hormonal risk factors, rare pathogenic variants in moderate and high risk breast/ovarian cancer susceptibility genes, and a polygenic risk score calculated using >300 single-nucleotide variants that explain ~20% of breast cancer polygenic variance. At present, BOADICEA has only been validated for women of European ancestry and might not be as accurate in other populations. The baseline (population) incidence is chosen according to the report by the Centers for Disease Control and Prevention and National Cancer Institute in 2020.

Coronary heart disease: The Pooled Cohort Equation (PCE) is an established tool used to estimate 10-year risk of coronary heart disease events, such as myocardial infarction, using demographic and clinical risk factors. These factors include age, sex, race, total cholesterol, HDL cholesterol, systolic blood pressure, hypertension treatment, current smoking status, diabetes diagnosis (PCE cannot be estimated if any of these factors are missing). The PCE was combined with polygenic risk to generate an integrated score for CHD. The PCE is validated in those aged 40+ and in White and African American populations. The PCE is not valid for those under the age of 40 and may not be as accurate in other populations. Risk associated with monogenic familial hypercholesterolemia (*LDLR, APOB, PCSK9,* and *LDLRAP1*) and family history of CHD are not accounted for in the PCE risk estimate. For more information on the PCE, please see reference: https://www.jacc.org/doi/pdf/10.1016/j.jacc.2013.11.005

**GIRA Generation:**
Determination of high risk for each condition evaluated was disease specific. See the tables below for the criteria used to determine high risk for each condition:

Table 1: Conditions evaluated in adult participants aged 18+ at time of enrollment.

| Condition | Age range assessed | PRS used to calculate risk | Genes Sequenced | Family History used to determine high risk | Integrated score using clinical data and PRS |
|---|---|---|---|---|---|
| Atrial fibrillation | 18+ | Yes | *LMNA* | Yes | No |
| Breast cancer | 18+ | Yes | *BRCA1, BRCA2, PALB2, PTEN, TP53, STK11* | Yes | BOADICEA (see methods) |
| Chronic kidney disease | 18+ | Yes | None | Yes | No |
| Colorectal cancer | 18+ | No | *EPCAM, MLH1, MSH2, MSH6, PMS2, STK11, PTEN, TP53* | No | No |
| Coronary heart disease | 18+ | Yes | *APOB, LDLR, LDLRAP1, PCSK9* | Yes | Pooled cohort equation (see methods) |
| Hypercholesterolemia | 18+ | Yes | *APOB, LDLR, LDLRAP1, PCSK9* | No | No |
| Obesity/BMI | 3+ | Yes | None | No | No |
| Prostate cancer | 18+ | Yes | *BRCA1, BRCA2, EPCAM, MLH1, MSH2, MSH6, PMS2* | Yes | No |
| Type 2 diabetes | 3+ | Yes | None | No | No |

Table 2: Conditions evaluated in pediatric participants aged 3-17 at time of enrollment.

| Condition | Age range assessed | PRS used to calculate risk | Genes Sequenced | Family History used to determine high risk | Integrated score using clinical data and PRS |
|---|---|---|---|---|---|
| Asthma | 3-17 | Yes | None | No | No |
| Obesity/BMI | 3+ | Yes | None | No | No |
| Type 1 diabetes | 3-17 | Yes | None | No | No |
| Type 2 diabetes | 3+ | Yes | None | No | No |

#### Limitations

Genetic research studies need large numbers of participants to understand how human DNA (or genes) contributes to disease risk. When research studies have low representation of some races, ethnicities, or ancestries (populations of descent), there is less genetic information available to understand risks for people in those groups. The GIRA health risk report has been validated (or confirmed) in up to four populations: Asian descent, African descent, European descent, or Hispanic/Latino descent. The report will name the populations included in the validation process. The estimate of risk may not be as accurate for some conditions if the participant is from a population that was not included in the validation process.

##### LIMITATIONS OF POLYGENIC RISK SCORES

Polygenic risk scores do not consider an individual's non-genetic factors such as lifestyle habits and personal or family medical history, which could affect risk. This research study used polygenic risk scores that were derived from people from several different populations. However, this type of information was not always available for every population. Although polygenic risk scores may be associated with disease risk in all populations, the scores are generally more accurate in people of European descent.

##### LIMITATIONS OF GENE SEQUENCING

All genetic tests have limitations. Some types of genetic variants may not be found by the gene sequencing test performed by Invitae. This means the test may rarely give an inaccurate result. If other family members are also tested, the Invitae test may find that family relationships are not what the participant believes them to be. Invitae will only report these findings if necessary to provide correct testresults.

### This is an example of the Invitae PDF.

The information in this file is sent through an API from invitae and matches the results that are in the json and parsed into the results field that feed into the summary

### eMERGE Polygenic Risk Report

| PATIENT INFORMATION | | REFERRING PROVIDER | | SPECIMEN | |
|---|---|---|---|---|---|
| Patient Name: | Malcolm Elliott | Provider Name: | Iam Testing | Report Type: | Final |
| Date of Birth: | 05/04/1977 | Referring Facility: | Cincinnati Children's Hospital Medical Center | Collected: | 05/15/2022 |
| Sample ID: | P8110050 | | | Received: | 01/01/2022 |
| Patient ID: | L81419710264 | | | Report Date: | 06/17/2022 |
| Accession ID: | SM-MO1BG | | | Material Type: | DNA |
| Site Sample ID: | N/A | Test Performed: | Polygenic Risk Evaluation | Material Source: | Saliva |
| | | Indication: | N/A | | |

#### Results Summary

In this patient the Polygenic Risk Score for the following condition(s) was determined to be **HIGH***:

coronary heart disease

*See detailed results for a description for how this risk was determined.

#### Detailed Results

This patient met the threshold for HIGH POLYGENIC RISK for the following condition(s):

##### Condition: coronary heart disease

1. A high polygenic risk score for coronary heart disease was found in this individual. A high polygenic risk score is associated with 1.7-2.3 times increased risk for developing coronary heart disease relative to a person not in the high risk category. The data is based on populations of European, African, Hispanic/Latino and Asian descent. Information is insufficient or not available for populations of other descent.
2. Factors including monogenic disease risk, family history, and other clinical measures can have an impact on the individuals overall (absolute) risk and should be considered.
3. This participant was tested as part of the Electronic Medical Records and Genomics (eMERGE) Genomic Risk Assessment and Management Study. The participant's integrated Genome Informed Risk Assessment (GIRA) report will be generated which will incorporate the results from this report as well as family history and monogenic risk status, if available.

#### Other Results

**Specific polygenic risk score details for coronary heart disease and breast cancer**

The specific polygenic risk score value (expressed as a z-score) for coronary heart disease and breast cancer are being reported for inclusion in absolute risk models that a healthcare provider may choose to use.

| Condition: | Test z-score: |
| --- | --- |
| breast cancer | N/A |
| coronary heart disease | 2.29 |

#### Limitations

- A polygenic score is neither deterministic nor diagnostic. Some people with a 'high risk' polygenic score will never develop the disease while others with a 'not high risk' polygenic score still have a risk of developing the disease that is equal to the general population. Therefore, this test is not intended to diagnose a disease or to make surgical or pharmacological intervention decisions. This test does not tell a patient anything about their current state of health and should not substitute for regular visits to the doctor. Any diagnostic or treatment decisions should be based on additional testing and/or other information that is managed by a healthcare provider.
- This test will detect genetic variants that are predefined. This test does not evaluate or report on all possible genetic variation related to a given disease. It will not detect novel or rare genetic variants and will not rule out the presence of these additional genetic variants related to a disease.
- The predefined list of genetic variants tested in this assay may be different from another institution or company, therefore genetic risk calculations and polygenic scores may differ if compared across different institutions or companies.
- The odds ratio (OR) listed for each condition does not take into account other factors that may play a role in a patient's overall risk of developing a disease (e.g family history or monogenic risk of developing the disease, or environmental and lifestyle risk factors).
- Receiving genetic test results may induce patient anxiety. Patients should speak to their doctor or healthcare provider regarding these test results and potential implications for their health and lifestyle decisions. If you have questions regarding your test results, you can contact the eMERGE clinical study team using the instructions provided to you during the consent process.
- Although the polygenic score has been developed to maximize the ability to predict risk in all ancestries, the availability of population reference data means that the score is currently most accurate for those with European ancestry. Due to this population limitation, as well as assay performance and processing issues, some patients may not receive a polygenic risk calculation for every condition listed. A result of "Not Resulted" for one condition does not impact the reliability of the risk polygenic score of other conditions.

#### Methodology

A genotyping microarray (the Global Diversity Array from Illumina, Inc.) was used to call single nucleotide variants (SNVs) at ~1.8 million sites in the genome. Only arrays meeting the QC criteria of >98% Call Rate were passed through to the downstream steps. Millions more SNVs in the sample were determined through imputation using data from the 1000 Genomes Project as a reference panel (Khera et al. 2018). Statistical association of SNVs and each condition listed were previously determined through examination of thousands of patient records and genomic data. The relative contribution of the set of SNVs associated with each condition were combined to provide a score. The score was further adjusted to account for the frequency of SNVs in different ancestry populations. The adjusted score was then represented as odds ratio (OR) and 95% confidence interval (CI) compared to a reference population.

The test is validated to determine polygenic risk for 10 conditions: asthma, atrial fibrillation, breast cancer, chronic kidney disease, coronary heart disease, hypercholesterolemia, obesity, prostate cancer, type 1 diabetes, and type 2 diabetes. Only conditions that meet the condition-specific criteria for high polygenic risk are reported as such by this test. In patients younger than 18 years of age, ONLY the following conditions are examined for risk: asthma, obesity, type 1 diabetes, and type 2 diabetes. Breast cancer is only assessed for individuals who elected their sex at birth as female and prostate cancer is only assessed for individuals who have elected their sex at birth as male. The methodologies used to determine the risk criteria for each condition are listed below:

***asthma*** polygenic risk status was determined based on a method that scores 985,837 sites in the genome. A Bayesian regression framework method was applied using the Trans-National Asthma Genetic Consortium (TAGC) GWAS to derive a multi-ancestral PRS score (PMID: 29273806). In a multiethnic study, individuals in the top 5% of the risk percentile had an increased risk of developing asthma. Values within the top 5% of this polygenic risk score are associated with a 1.95 odds ratio (OR) in pediatric populations of European descent at a 95% CI [1.43-2.65], 1.83 OR in pediatric populations of African descent at a 95% CI [1.24-2.70], and 3.12 OR in pediatric populations of Hispanic/Latino descent at 95% CI [1.32-7.44]. Information is insufficient or not available for populations of other descent.

***atrial fibrillation*** polygenic risk status was determined based on a method developed by Nielsen et al. (PMID: 30061737) that scores 161 sites in the genome. In a multiethnic study, individuals in the top 3% of the risk percentile exhibited an increased risk of developing atrial fibrillation. Values within the top 3% of this polygenic risk score are associated with a 2.32 OR in populations of European descent at a 95% CI [2.07-2.61], 2.19 OR in populations of African descent at a 95% CI [1.38-3.38], and 2.27 OR in populations of Hispanic/Latino descent at a 95% CI [1.09-4.50]. Information is insufficient or not available for populations of other descent.

***breast cancer*** polygenic risk status was determined based on a method developed by Mavaddat et al. (PMID: 30554720) that scores 308 sites in the genome. In a multiethnic study, individuals who fell in the top 5% of the risk percentile exhibited an increased risk of developing breast cancer. Values within the top 5% of this polygenic risk score are associated with a 2.47 OR in populations of European descent at a 95% CI [2.20 - 2.77] (PMID: 30554720), 1.61 OR in populations of African descent at a 95% CI [1.38-1.87] (PMID: 33769540), 2.05 OR in populations of Hispanic/Latino descent at a 95% CI [1.10-3.83] (PMID: 34347061), and 2.22 OR in populations of Asian descent at a 95% CI [1.99-2.47] (PMID: 32737321). Information is insufficient or not available for populations of other descent.

***chronic kidney disease*** polygenic risk status was determined based on a study of renal function by Wuttke et al. (PMID: 31152163) and includes 471,316 sites in the genome. In a multiethnic validation study, individuals who fell in the top 2% of the risk percentile exhibited an increased risk of developing chronic kidney disease. Values within the top 2% of this polygenic risk score are associated with a 3.60 OR in populations of European descent at a 95% CI [3.11-4.17], 2.66 OR in populations of African descent at a 95% CI [2.01-3.51], 4.93 OR in populations of Hispanic/Latino descent at a 95% CI [2.46-9.89], and 2.81 OR in populations of Asian descent at a 95% CI [1.91-7.59]. Information is insufficient or not available for populations of other descent.

***coronary heart disease*** polygenic risk status was determined based on a method that scores 458,384 sites in the genome. In a multiethnic study, individuals in the top 5% of the risk percentile exhibited an increased risk of developing coronary heart disease. Values within the top 5% of this polygenic risk score are associated with a relative risk of 2.30 in populations of European descent at a 95% CI [2.07-2.56], 1.68 in populations of African descent at a 95% CI 1.39-2.032], and 2.16 in populations of Hispanic/Latino descent at a 95% CI [1.47-3.19]. For Asian populations, the relative risk is similar to that in European populations. Information is insufficient or not available for populations of other descent.

***hypercholesterolemia*** polygenic risk status was determined based on a method that scores 9,009 sites in the genome. In a multiethnic study, individuals who fell in the top 3% of the risk percentile exhibited an increased risk of developing hypercholesterolemia. Values within the top 3% of this polygenic risk score are associated with a 4.16 OR in populations of European descent at a 95% CI [2.59-6.44], 3.16 OR in populations of African descent at a 95% CI [1.92-5.01], 4.02 OR in populations of Hispanic/Latino descent at a 95% CI [2.72-5.83], and 3.75 OR in populations of Asian descent at a 95% CI [3.15-4.42]. Information is insufficient or not available for populations of other descent.

***obesity*** polygenic risk status was determined based on a method that scores 1,217,710 sites in the genome. In a multiethnic study, individuals who fell in the top 3% of the risk percentile exhibited an increased risk above baseline of developing obesity. Values within the top 3% of this polygenic risk score are associated with a 4.08 OR in populations of European descent at a 95% CI [3.02-5.52], 2.54 OR in populations of African descent at a 95% CI [1.66-3.98], 2.33 OR in populations of Hispanic/Latino descent at a 95% CI [1.64-3.31], and 5.73 OR in populations of Asian descent at a 95% CI [2.28-14.57]. Information is insufficient or not available for populations of other descent.

***prostate cancer*** polygenic risk status was determined based on a method developed by Conti et al. (PMID: 33398198) that scores 264 sites in the genome. In this multiethnic study of individuals of African, European, Asian, and Hispanic/Latino descent, individuals who fell in the top 10% of the risk percentile exhibited an increased risk of developing prostate cancer. Values within the top 10% of this polygenic risk score are associated with a 3.67 OR in populations of European descent at a 95% CI [3.57-3.76] and a 2.95 OR in populations of African descent at a 95% CI [2.60-3.30]. Information is insufficient or not available for populations of other descent.

***type 1 diabetes*** polygenic risk status was determined based on a method developed by Sharp et al. (PMID: 30655379) that scores 71 sites in the genome. In a separate multiethnic study, individuals who fell in the top 3% of the risk percentile exhibited an increased risk of developing type 1 diabetes. Values within the top 3% of this polygenic risk score are associated with a 12.97 OR in populations of European descent at a 95% CI [7.29-20.40] and 20.45 OR in populations of African descent at a 95% CI [10.77-38.83]. Information is insufficient or not available for populations of other descent.

***type 2 diabetes*** polygenic risk status was determined based on a method developed by Ge et al. (medRxiv 2021.09.11.21263413) that scores 1,259,754 sites in the genome. In a multiethnic study, individuals who fell in the top 2% of the risk percentile exhibited an increased risk of developing type 2 diabetes. Values within the top 2% of this polygenic risk score are associated with a 4.21 OR in populations of European descent at a 95% CI [3.66-4.84], 2.55 OR in populations of African descent at a 95% CI [2.09-3.11], 4.58 OR in populations of Asian descent at a 95% CI [4.00-5.23], and 6.87 OR for Hispanic/Latino populations at a 95% CI [3.11, 15.15]. Information is insufficient or not available for populations of other descent.

**Testing performed under the direction of Heidi L. Rehm, PhD, FACMG**

**Results reviewed and approved for release by:** Heidi L Rehm **06/17/2022**

### MeTree Family Health History

**Patient Name:** Jane Smith
**Date of Birth:** 01/01/1984

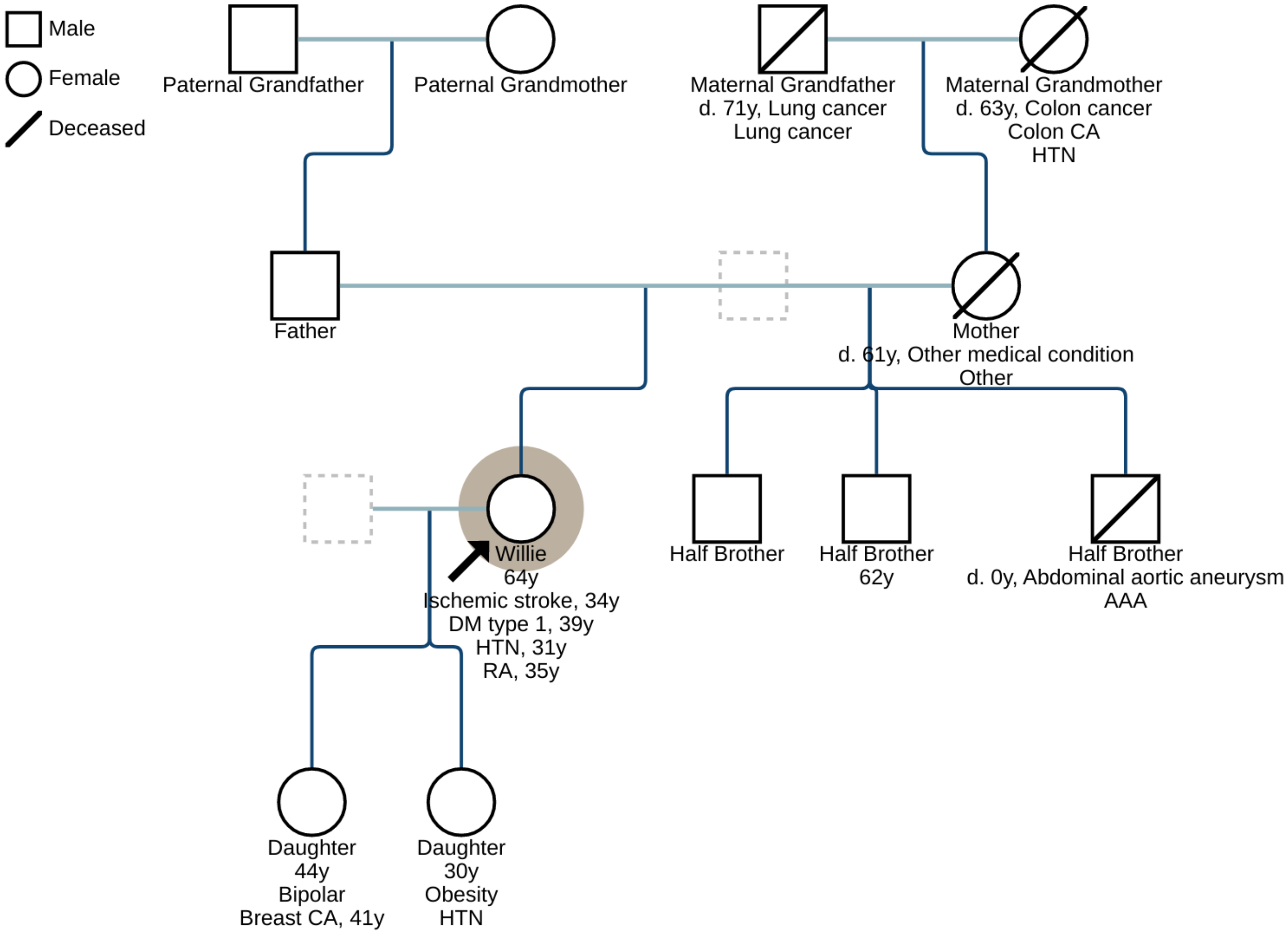
