## Supplemental 2 for "Implementing Integrated Genomic Risk Assessments for Breast Cancer: Lessons Learned from the eMERGE Study"

| | |
|---|---|
| Record ID | 115673 |
| GIRA Report Date | 09-21-2023 |

#### eMERGE Study Overview

eMERGE is a research network of 10 academic medical centers across the United States. It is funded by the National Human Genome Research Institute (NHGRI). The eMERGE study aims to find better ways to assess and manage patients' risk for future health conditions. eMERGE wants to learn if risk based on genetics, family health history, and personal health history, helps patients and their doctors make health care choices. This study is developing a Genome Informed Risk Assessment (GIRA) report. The GIRA report will have information about genetic risk, clinical risk, and family history for certain common conditions in adults and children.

#### Sections included in the GIRA Report

- Summary of findings
- Risk result breakdown
- Patient education page(s)
- Participant frequently asked questions
- Study methods and limitations
- Invitae eMERGE panel screen report
- Broad Institute Polygenic Risk Report
- MeTree family health history pedigree

#### Site Contacts


| | |
|---|---|
| Vanderbilt University Medical Center<br>Principal Investigator<br>Dan M. Roden, MD<br>phone: 615-322-0067<br> | Vanderbilt University Medical Center<br>Study Coordinator<br>Harris Bland, MPH, MBA<br>phone: 615-875-8240<br> |


#### Genome Informed Risk Assessment (GIRA)

| | | |
|---|---|---|
| The following report is a summary of evaluated risk factors for common conditions. Common gene changes were tested for the conditions listed. Rare gene changes and family history were also included for some conditions. This study determined who is at high risk based on specific criteria for each condition. | Risks Evaluated: | |
| | Asthma* | Hypercholesterolemia |
| | Atrial fibrillation | Obesity/BMI** |
| | Breast cancer | Prostate cancer |
| | Chronic kidney disease | Type 1 diabetes* |
| | Colorectal cancer | Type 2 diabetes** |
| | Coronary heart disease | |

* children only **adults and children

| eMerge network ELECTRONIC MEDICAL RECORDS AND GENOMICS | **Name:** Jane Smith | **Provider:** John Tester |
|---|---|---|
| | **DOB:** 1977-05-02 | **Site ID:** Vanderbilt University Medical Center |
| | **Sex at birth:** Female | **Participant Lab ID:** L712345677 |
| | **Date of Report:** 09-21-2023 | |

#### Summary of Findings

**RESULT: This individual was found to be at high risk for one or more of the conditions evaluated.**

Being at high risk for a condition does not mean that this individual will definitely get that condition. When high risk for a condition is identified, recommendations are provided that may help reduce the risk of getting the condition or help treat the condition.

**For participants already diagnosed with a condition, providers should continue with current treatment.**

#### This participant is at high risk for the following:

• Breast Cancer (Integrated Score)

*****Breast Cancer*****
**----------------------------------------------------------------------------------------------------**

Risk Category: **Integrated Score**

**Care Recommendations:**
• Emphasize a healthy lifestyle:
 – Exercise regularly.
 – Maintain healthy body weight.
 – Diet rich in fruit and vegetables.
 – Limit alcohol intake.
• Consider consultation with a breast specialist about medications to reduce the risk of breast cancer (tamoxifen or other anti-estrogen medications).
• Consider risks and benefits of hormone replacement therapy.
• Consider annual mammogram starting at age 40 or 10 years before the youngest breast cancer in the family, whichever is younger.
• Consider annual breast MRI starting at age 40 or 10 years before the youngest breast cancer in the family, whichever is younger.

**Professional Guidelines:**
• Breast cancer NCCN Guidelines

For any condition studied that is not listed in the table above, high risk was not identified. This means the polygenic risk scores (PRS) for these conditions did not meet the threshold for high risk. For some conditions, other factors such as sequencing of specific genes (monogenic risk), or family history, also contributed to the risk estimate.

Review the methodologies section for a full explanation of how high risk for each condition was determined.

**Please note that a person's overall risk for any of these conditions could still be higher than the general population based on factors that are not included in the GIRA.**

Information contained in this report does not replace evaluation by a health care provider. General risk reducing strategies such as maintaining a healthy lifestyle and age recommended screening tests are still recommended.

Questions about what this report means for this individual's medical management should be discussed with their provider.

| | | |
|---|---|---|
| eMerge network ELECTRONIC MEDICAL RECORDS AND GENOMICS | **Name:** Jane Smith | **Provider:** John Tester |
| | **DOB:** 1977-05-02 | **Site ID:** Vanderbilt University Medical Center |
| | **Sex at birth:** Female | **Participant Lab ID:** L712345677 |
| | **Date of Report:** 09-21-2023 | |

| **Risk Result:** | **High Risk for Breast Cancer** |
|---|---|

#### Monogenic Results: Negative

Gene sequencing did not identify any pathogenic or Likely Pathogenic variants related to this condition. See the Invitae report for a full list of genes sequenced.

#### Integrated Risk Score: High Risk

Based upon the participant's genetic risk, family history, and personal factors including age, sex, body mass index, breast density, reproductive history, and hormonal exposure, the lifetime risk of breast cancer to the age of 80 is 37.6%. The average woman has a lifetime risk of breast cancer of 12%, so this risk is higher.

**Limitations of polygenic risk:**

This polygenic risk does not take into account the individual's non-genetic factors such as lifestyle, habits and history of other diseases, which could affect risk. These results should be viewed in the context of the individual's medical care, family history, and racial/ethnic background. See the full methods and limitations for additional information.

### Breast Cancer: Understanding Your Results

#### What is breast cancer?

- Cancer is a disease in which cells in the body grow out of control. When cancer starts in the breast, it is called breast cancer. Although many types of breast cancer can cause a lump in the breast, not all do. Breast cancer can spread when the cancer cells get into the blood or lymph system and are carried to other parts of the body.
- Risk factors for breast cancer include variants in your genes, having other family members with breast cancer (family history), and your age. Your reproductive history can affect your risk for breast cancer. This includes how many children you have had, whether you breast-fed your children, and when you started and stopped having your period. There are also lifestyle factors that affect an individual's risk for breast cancer including use of hormonal birth control, estrogen replacement therapy, weight and how much alcohol you drink.
- You can learn more about breast cancer and risk factors here (https://www.cancer.org/cancer/breast-cancer/about.html)

#### What does high risk for breast cancer mean?

- On average, 12-13 out of 100 women, about 12-13% of women, will get breast cancer in their lifetime, by the age of 80 years.
- High risk for breast cancer means that you have a risk of getting breast cancer of 25 or more out of 100, or 25% or greater in your lifetime, by the age 80 years. *Please see your test report for more information about your specific lifetime risk for breast cancer.*

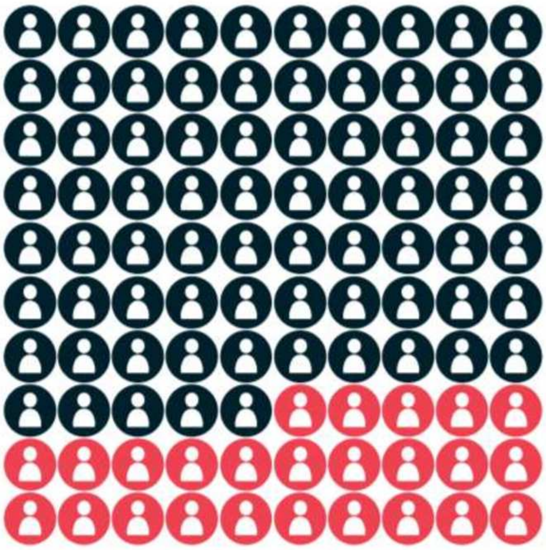

This picture shows high risk for breast cancer. 25 women* (red) get breast cancer and 75 women (gray) do not.

This picture shows overall high risk for breast cancer. *Your risk may be the same as this or higher.* Please look at your test report for your specific risk.

*Women here refers to individuals assigned a female sex at birth.

- The breast cancer risk score is integrated, meaning it takes multiple sources of risk into account. The integrated risk score includes genetic factors such as polygenic risk and family history and non-genetic factors such as reproductive history, medicine that affect hormone levels, weight and age. This integrated score included genetic information from large research studies of people with European, East Asian, African, and Hispanic/Latino descent.

#### What can you do to lower your risk or detect breast cancer early?

- Not everyone who is at high risk for breast cancer will get it.
- Early breast cancer detection:
  - Continue or start to perform monthly breast self-exams ((https://www.breastcancer.org/symptoms/testing/types/self_exam). This is important to help you know your own breasts and notice any changes as soon as possible. You should talk to your doctor if you notice any changes.
  - For women at high risk for breast cancer, we suggest alternating mammogram and breast MRI every 6 months, beginning at age 40 or 10 years younger than the youngest family member's breast cancer diagnosis, whichever is younger.
- To lower your risk:
  - Talk to your doctor about estrogen-blocking medications like tamoxifen.
  - Talk to your doctor about avoiding medications containing estrogen.
  - Consider breast feeding if appropriate.
  - Maintain a healthy body weight with a body mass index (BMI) (https://www.cdc.gov/healthyweight/assessing/bmi/adult_bmi/english_bmi_calculator/bmi_calculator.html) < 25, eat a healthy diet, keep physically active and limit alcohol intake.
  - Surgical removal of healthy breasts is typically not recommended, but discuss this with your doctor if you have concerns.

#### What are your next steps?

- You should share these results with your doctor(s) or other healthcare provider to discuss actions to lower your risk.
- You may also want to share your results with your family members.
- Your results will be uploaded to your electronic health record for you to review and will be available to your doctor(s) and other healthcare providers.
- If you have any questions about your results, please contact the study team at your institution. You can find this contact information on the cover page of the GIRA.

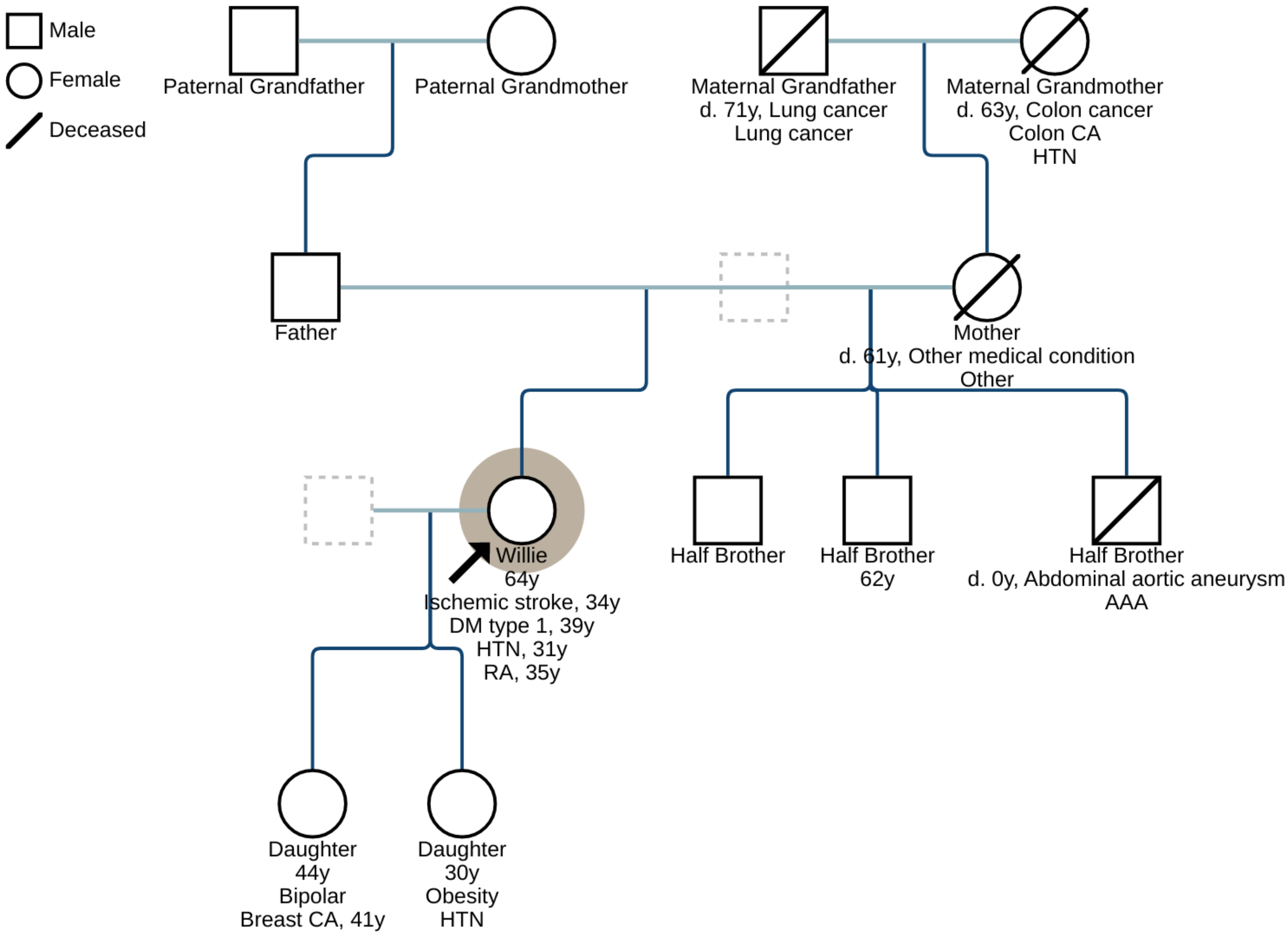
