## Supplemental 3 for "Implementing Integrated Genomic Risk Assessments for Breast Cancer: Lessons Learned from the eMERGE Study"

**For participants already diagnosed with a condition, providers should continue with current treatment.**

### This participant is at high risk for the following:

• BRCA1 (Monogenic Risk)

*****Gene: BRCA1*****

**-----------------------------------------------------------------------------------------------------**

Risk Category: **Monogenic Risk**

**Associated Risks:**
• Increased risk for breast, ovarian, prostate, pancreatic, and possibly melanoma cancers.
• See risk results page and attached Invitae report for additional information.

**Care Recommendations:**
• If your patient has not yet spoken to a genetic counselor regarding their results, we recommend referring your patient to a genetic counselor to discuss their results and potential next steps.
• See nccn.org for management guidelines of individuals with pathogenic variants in this gene.
• Please refer to the Invitae report and Invitae positive results guide for guidance.

**Professional Guidelines:**
• Breast cancer NCCN Guidelines

For any condition studied that is not listed in the table above, high risk was not identified. This means the polygenic risk scores (PRS) for these conditions did not meet the threshold for high risk. For some conditions, other factors such as sequencing of specific genes (monogenic risk), or family history, also contributed to the risk estimate.

Review the methodologies section for a full explanation of how high risk for each condition was determined.

**Please note that a person's overall risk for any of these conditions could still be higher than the general population based on factors that are not included in the GIRA.**

Information contained in this report does not replace evaluation by a health care provider. General risk reducing strategies such as maintaining a healthy lifestyle and age recommended screening tests are still recommended.

Questions about what this report means for this individual's medical management should be discussed with their provider.

| eMerge network ELECTRONIC MEDICAL RECORDS AND GENOMICS | **Name:** Jane Smith | **Provider:** John Tester |
|---|---|---|
| | **DOB:** 1984-01-01 | **Site ID:** Vanderbilt University Medical Center |
| | **Sex at birth:** Female | **Participant Lab ID:** L712345677 |
| | **Date of Report:** 03-25-2025 | |

| **Risk Result:** | **High Risk for Breast Cancer** |
|---|---|

### Monogenic Results: Positive

A pathogenic or Likely Pathogenic variant associated with hereditary breast and ovarian cancer was identified in the BRCA1 gene. People with a pathogenic or Likely Pathogenic variant in BRCA1 are at increased risk for breast, ovarian, prostate, pancreatic, and possibly melanoma cancers. See attached Invitae report for additional risk information.

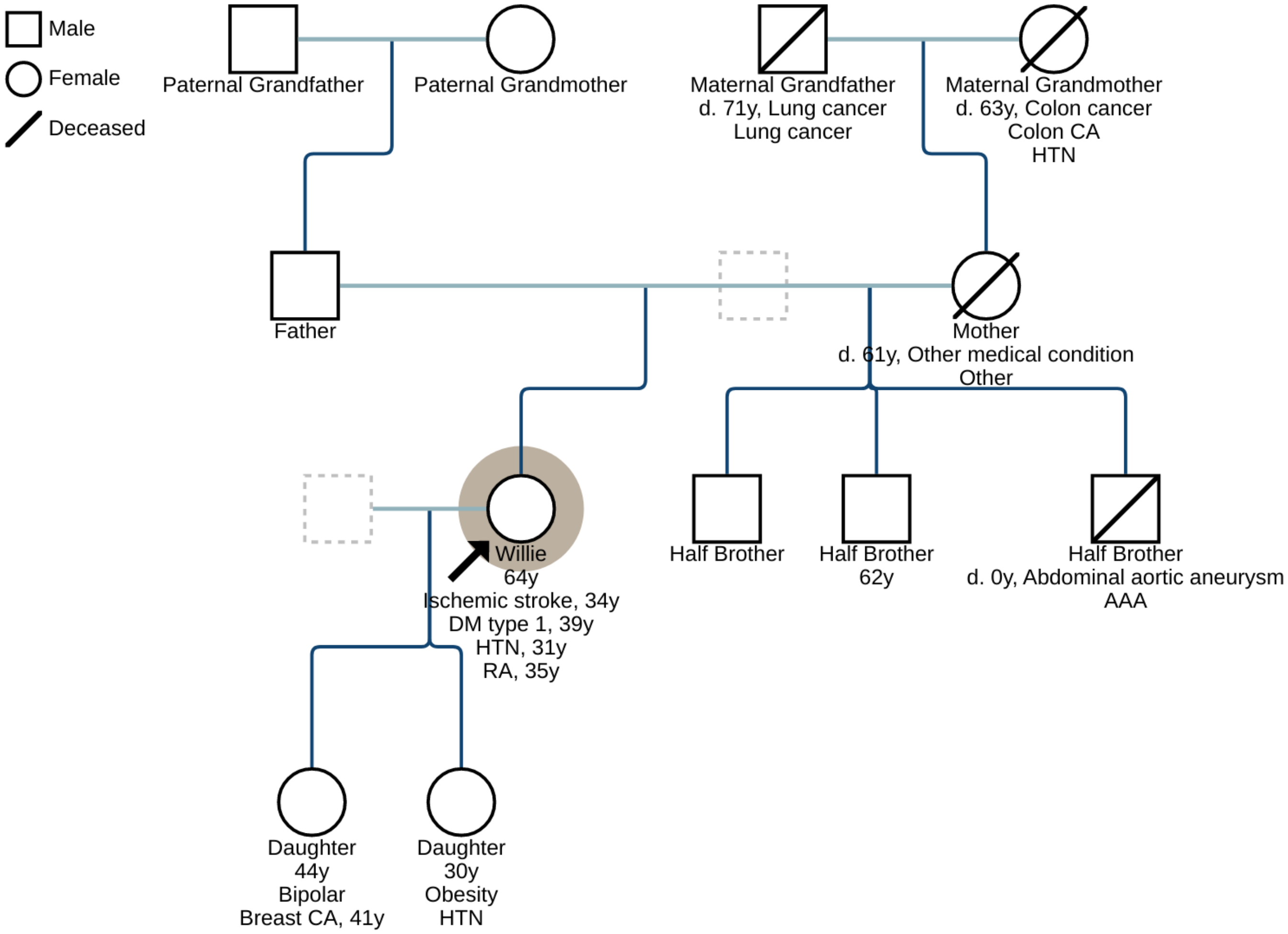
